# Ubie Symptom Checker: A Clinical Vignette Simulation Study

**DOI:** 10.1101/2024.08.29.24312810

**Authors:** N. Kenji Taylor, Takashi Nishibayashi

## Abstract

**Background:** AI-driven symptom checkers (SC) are increasingly adopted in healthcare for their potential to provide users with accessible and immediate preliminary health education. These tools, powered by advanced artificial intelligence algorithms, assist patients in quickly assessing their symptoms. Previous studies using clinical vignette approaches have evaluated SC accuracy, highlighting both strengths and areas for improvement.

**Objective:** This study aims to evaluate the performance of the Ubie Symptom Checker (Ubie SC) using an innovative large language model-assisted (LLM) simulation method.

**Methods:** The study employed a three-phase methodology: gathering 400 publicly available clinical vignettes, medical entity linking these vignettes to the Ubie SC using large language models and physician supervision, and evaluation of accuracy metrics. The analysis focused on 328 vignettes that were within the scope of the Ubie SC with accuracy measured by Top-5 hit rates.

**Results:** Ubie achieved a Top-5 hit accuracy of 63.4% and a Top-10 hit accuracy of 71.6%, indicating its effectiveness in providing relevant information based on symptom input. The system performed particularly well in domains such as the nervous system and respiratory conditions, though variability in accuracy was observed across different ICD groupings, highlighting areas for further refinement. When compared to physicians and comparator SC’s that used the same clinical vignettes set, Ubie compared favorably to the median physician hit accuracy.

**Conclusions:** The Ubie Symptom Checker shows considerable promise as a supportive education tool in healthcare. While the study highlights the system’s strengths, it also identifies areas for improvement suggesting continued refinement and real-world testing are essential to fully realize Ubie’s potential in AI-assisted healthcare.

## Introduction

The adoption of AI-driven symptom checkers in healthcare is rising due to their potential to provide users with accessible and immediate preliminary health education. These tools, powered by advanced artificial intelligence algorithms, have become valuable for patients seeking quick assessments of their symptoms [1]. Numerous studies have evaluated the accuracy of these symptom checkers, often using clinical vignette approaches. These studies highlight both the strengths and weaknesses of symptom checkers, providing valuable insights into their effectiveness and areas for improvement [2, 3, 4].

The Ubie Symptom Checker (SC), a service available in the US, is a notable example of such a tool. Utilizing a combination of publicly available data, engineering expertise, and large language models, the Ubie algorithm, in collaboration with physicians and patient feedback, maps symptoms to provide accurate assessments. Ubie SC not only compares its data against user feedback but also is constantly being updated by a team of physicians based on incoming data from patient and health provider network clinical feedback. To measure the performance of Ubie SC, we adopted a clinical vignette approach [5]. Clinical vignettes are widely used in the training and evaluation of healthcare providers (physicians, nurse practitioners, physician assistants) focused on diagnosis and treatment [6]. Consequently, clinicians evaluating these new technologies are familiar and comfortable with this methodology as a performance benchmarking tool. Vignette studies can be comprehensive and varied in scope while maintaining control over the testing environment to focus purely on the AI SC performance as a user information tool. Additionally, vignettes and their results are publicly available, making replication and comparison to other studies more straightforward.

While vignette studies have well-known benefits, a major downside is their static nature. Most vignette studies involve manual entry, including data extraction or interpretation phases, and manual input of set parameters followed by accuracy metrics. Consequently, they are often smaller in sample size, less comprehensive, and resource-intensive [7]. We sought to address these limitations by developing an AI-assisted simulation method utilizing publicly available validated clinical vignettes to:

1. Explore the use of AI-assisted simulation for SC accuracy
2. Understand Ubie’s SC accuracy based on clinical vignette data as it compares to other SC and physician accuracy

This innovative approach aims to overcome the static nature of traditional vignette studies, allowing for dynamic simulations that enable ongoing testing of the AI models and facilitate modifications to improve future accuracy.

## Methods

As shown in Figure 1, our methodology was systematically structured into 3 distinct phases: gathering clinical vignettes, mapping vignettes using a combination of large language model (LLM) assistance and physician supervision, and evaluating accuracy metrics.

**Figure 1.**
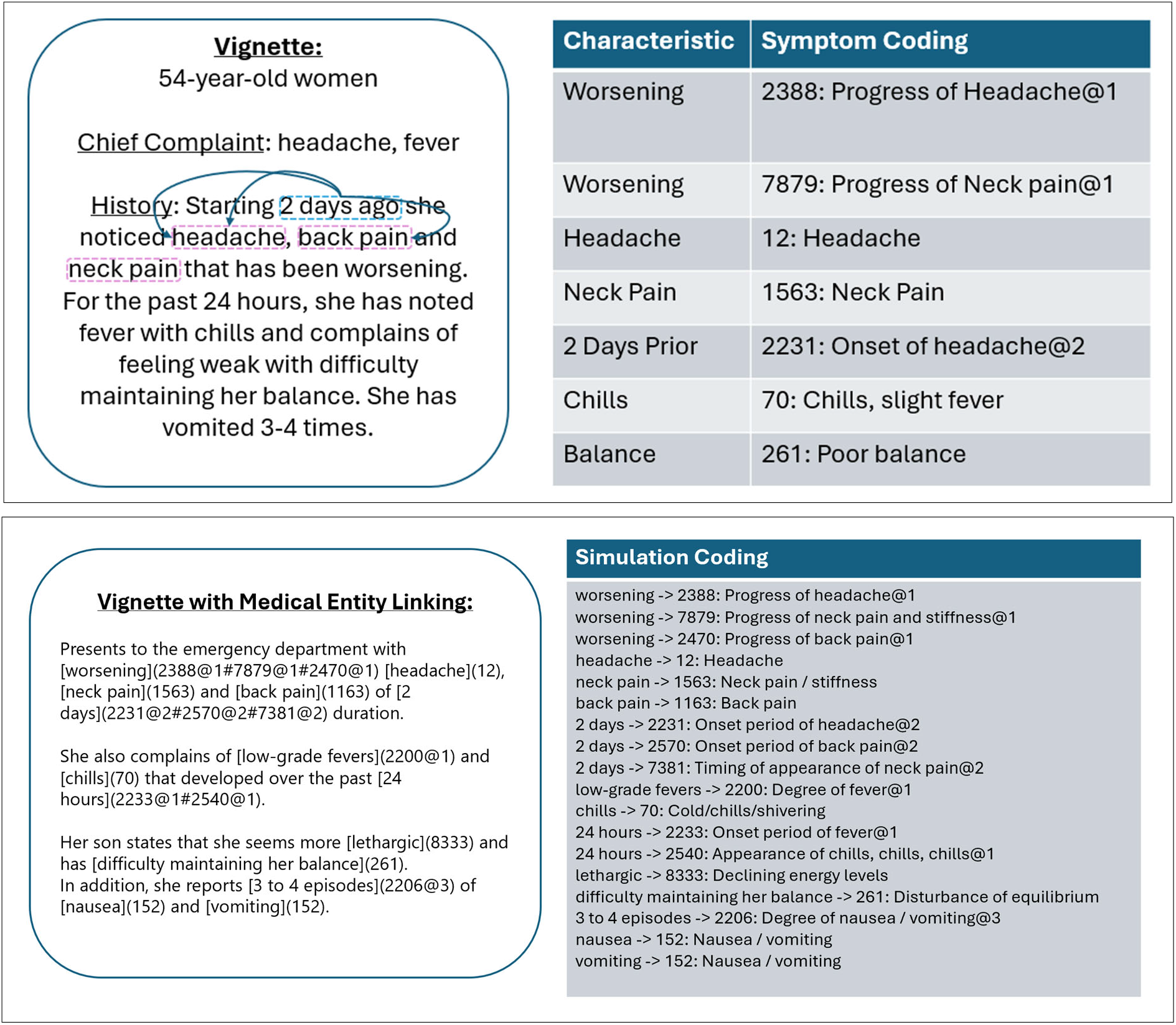
Medical Entity Linking Example.

### Phase 1: Gathering of Vignettes

As the foundation of our study, we utilized 400 publicly available, peer-reviewed clinical vignettes. These vignettes were rigorously created, tested, and validated following the recommended methodologies in the field[8]. Each vignette included a “gold standard” diagnosis, detailed case information, and a differential diagnosis. The vignettes were selected for their volume, breadth, and ease of access, ensuring a comprehensive dataset for our analysis. This robust selection process was aimed at providing a diverse and representative sample of real-world clinical scenarios.

### Phase 2: Mapping of Vignettes (Figure 1)

Medical information systems convert concepts such as diseases and symptoms into specific knowledge representations (e.g. ICD-10) for use in a variety of contexts from research to insurance reimbursement to clinical care. However, there is no uniform system of representation that is easy for SC’s to use. Hence, SC providers have their own symptom knowledge base. Since the Ubie SC also uses a unique system of representation, symptoms contained in the vignettes need to be converted through an iterative process known as medical entity linking [9, 10].

The components of each vignette used for mapping to the Ubie SC simulator included:

1. Case Number
2. Age
3. Sex
4. Chief Complaint(s)
5. Presentation
6. Absent Findings
7. Physical Exam Findings
8. Physical History
9. ICD-10 Code

Initially, LLM assistance provided a preliminary mapping to the Ubie SC coding system to simulate a user inputting the data directly into the SC platform. These codes were then refined and reviewed by AI engineer (TN) and physician (NKT). This dual-layered approach ensured both computational efficiency and clinical accuracy as mapping had to be completed across all components of the vignette. For example, a viral upper respiratory infection needed mapping to the corresponding SC code representing a viral upper respiratory infection. Similarly, each individual chief complaint needed a map to the corresponding SC code. The iterative mapping process is illustrated in Figure 1, which highlights the transition from initial mapping to simulation coding. The incorporation of two layers of supervision ensured that the mappings were clinically relevant and accurate, reducing the risk of errors that could arise from an automated process alone.

### Phase 3: Accuracy Analysis

To evaluate the accuracy of the symptom checker, we used Top-N as our primary metric. The Top-N accuracy is a commonly used metric in the research and commercial field. Instead of the full 400 vignettes, our primary focus was on the 328 vignettes for which the Ubie SC could provide diseases related to input symptoms. The exclusion of 75 vignettes was due to the absence of the exact diseases in the scope of the SC, as shown in the methodology diagram (Figure 2). For exa ple, brucellosis is not with the scope fo the Ubie SC database at the time of study so was not included as a test case because the SC was not designed to display this disease name related to symptoms input by the user.

**Figure 2.**
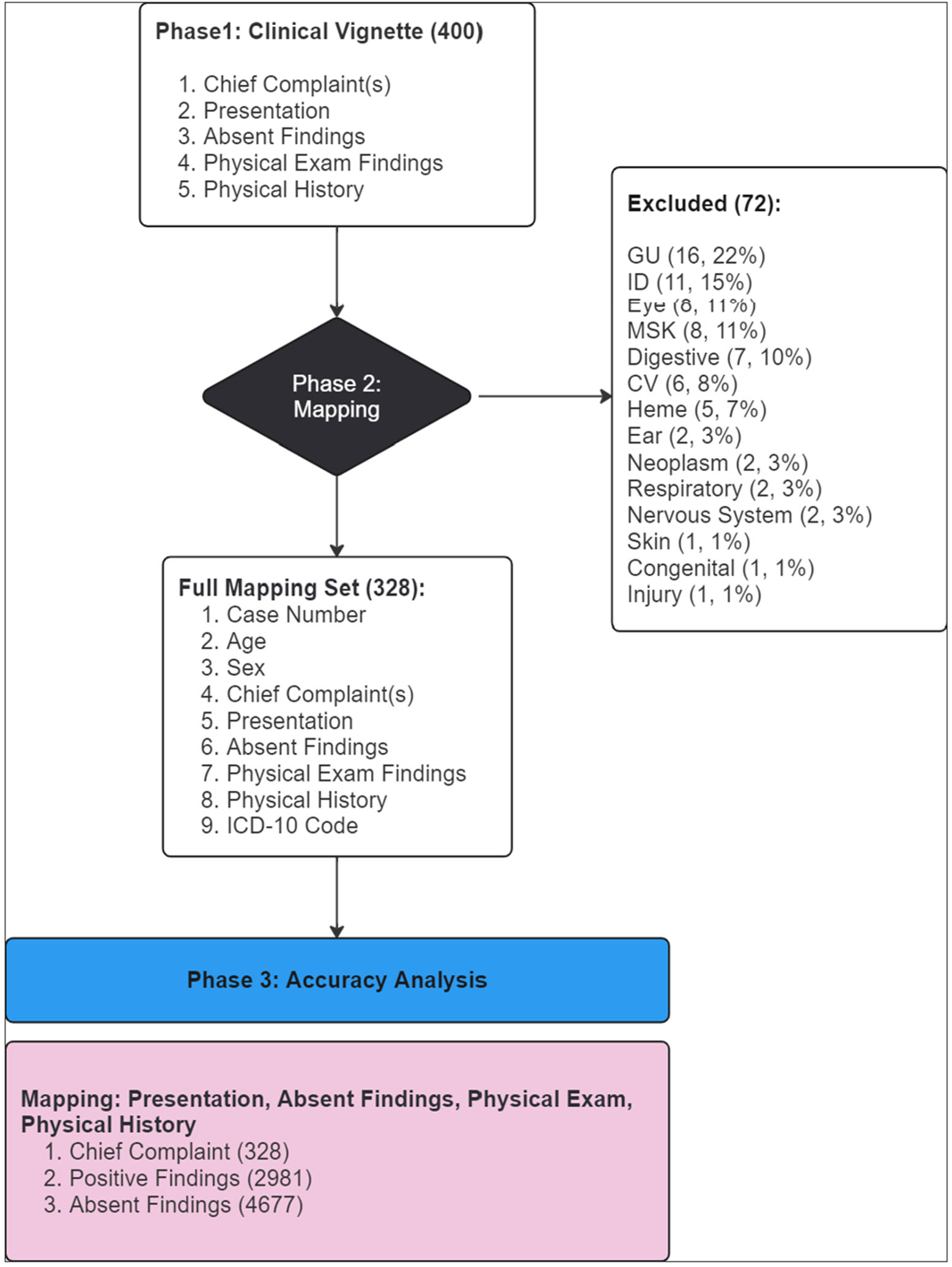
Vignette Characteristics. Numbers in parentheses represent the unique number of data elements simulated at each stage.

For accuracy measures, unless otherwise noted, the denominator was the number of vignettes where the gold standard disease was within the scope of the Ubie SC. This approach ensured that our evaluation was reflective of the SC’s intended scope of functionality.

## Results

### A. Vignette Characteristics (Figure 2)

Eight-two percent (328) out of the 400 original vignettes had gold standard diagnoses that were within scope of the SC (Figure 1). Of the 72 diseases that were out of scope within the SC, the majority fell under the categories of genitourinary (ie: overflow incontinence, urge incontinence, urogenital fistula), infectious disease (ie: brucellosis, cat scratch disease, cholera), eye conditions (ie: stye, chalazion) and musculoskeletal conditions (ie: sciatica, rotator cuff injury and achilles tendinitis).

From the 328 vignettes that were mapped, a total of 2981 (range 1 to 21) non-unique symptoms were mapped to the SC coding system for simulation. This represents a mean of 9.1 present symptoms (ie: fever, cough, tachycardia, right upper abdominal pain) or physical exam findings per vignette. There were a total of 4677 (range 3 to 26) non-unique absent symptoms (ie: no fever, no cough, no tachycardia) that were mapped to the SC simulator. The mean number of non-unique absent symptoms per vignette was 14.3.

The vignettes covered a broad range of conditions as noted in table 1. The majority were vignettes of people who were over the age of 18 (298, 90.9%) with slightly more cases involving male sex (176, 53.7%). Almost a third of vignettes (102, 31.0%) were repeated with a different presentation at least more than once. A diagnosis of COPD, leukemia, unstable angina, stable angina, heart failure and pulmonary embolism were the diagnoses with 4 different vignette presentations.

**Table 1.** Vignettes within Scope by ICD Grouping.

| ICD Grouping | Count | Percentage |
| --- | --- | --- |
| Infectious Disease | 49 | 14.9 |
| Circulatory system | 39 | 11.9 |
| Endocrine system | 32 | 9.8 |
| Genitourinary system | 32 | 9.8 |
| Neoplasm | 31 | 9.5 |
| Respiratory system | 28 | 8.5 |
| Digestive system | 25 | 7.6 |
| Musculoskeletal system | 21 | 6.4 |
| Obstetrics | 16 | 4.9 |
| Nervous system | 15 | 4.6 |
| Ear-related | 11 | 3.4 |
| Hematology | 9 | 2.7 |
| Ophthalmology | 7 | 2.1 |
| Dermatology | 5 | 1.5 |
| Injury, poisoning | 4 | 1.2 |
| Congenital malformations | 2 | 0.6 |
| Other | 2 | 0.6 |
| Total | 328 | 100.0 |

### B. Accuracy of SC across domains

The SC is programmed to provide a related disease based on symptoms of up to 10 diseases. A “hit” is defined as a match between the SC presentation and the gold standard diagnosis assigned to each vignette (Figure 3). Out of the total 328 vignettes, there were a total of 234 hits (71.6%). In other words, the SC produced the matching disease in the top 10 differential 71.6% of the time. The overall Top 1, 5 and 10 hit accuracy was 29.3%, 63.4% and 71.6% respectively. As shown in Figure 3, 40.9% of the hits came in the top 1 rank, 71.9% by the top 3 ranking, 88.5% in the Top 5 and 94.9% in the top 8. Top 5 hit accuracy is presented given the vast majority of hits fell within the Top 5 and is a commonly presented metric in similar evaluations of SCs in prior studies.

**Figure 3.**
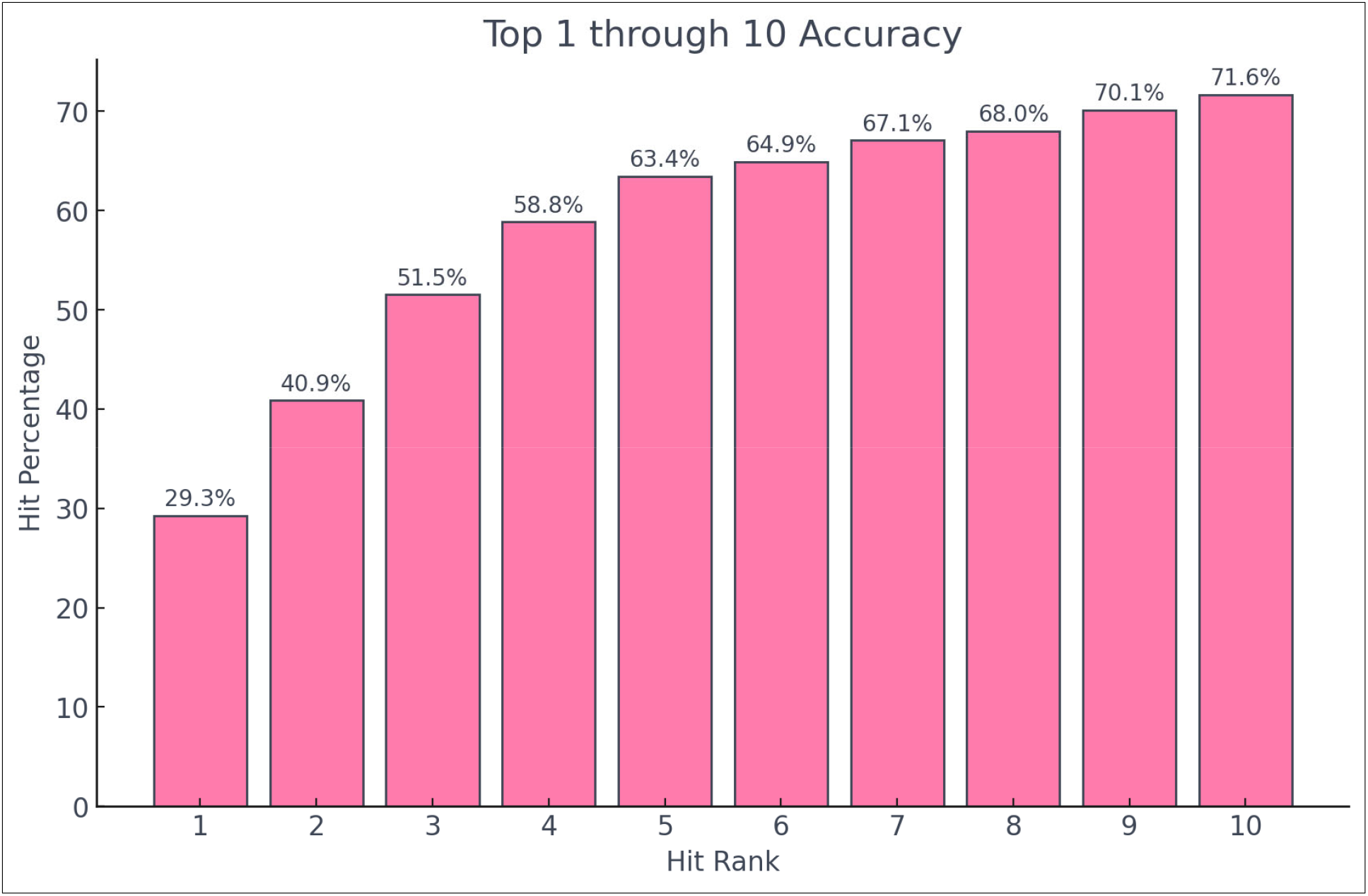
Cumulative Hits by Rank (Top 1 through 10)

**Figure 3.**
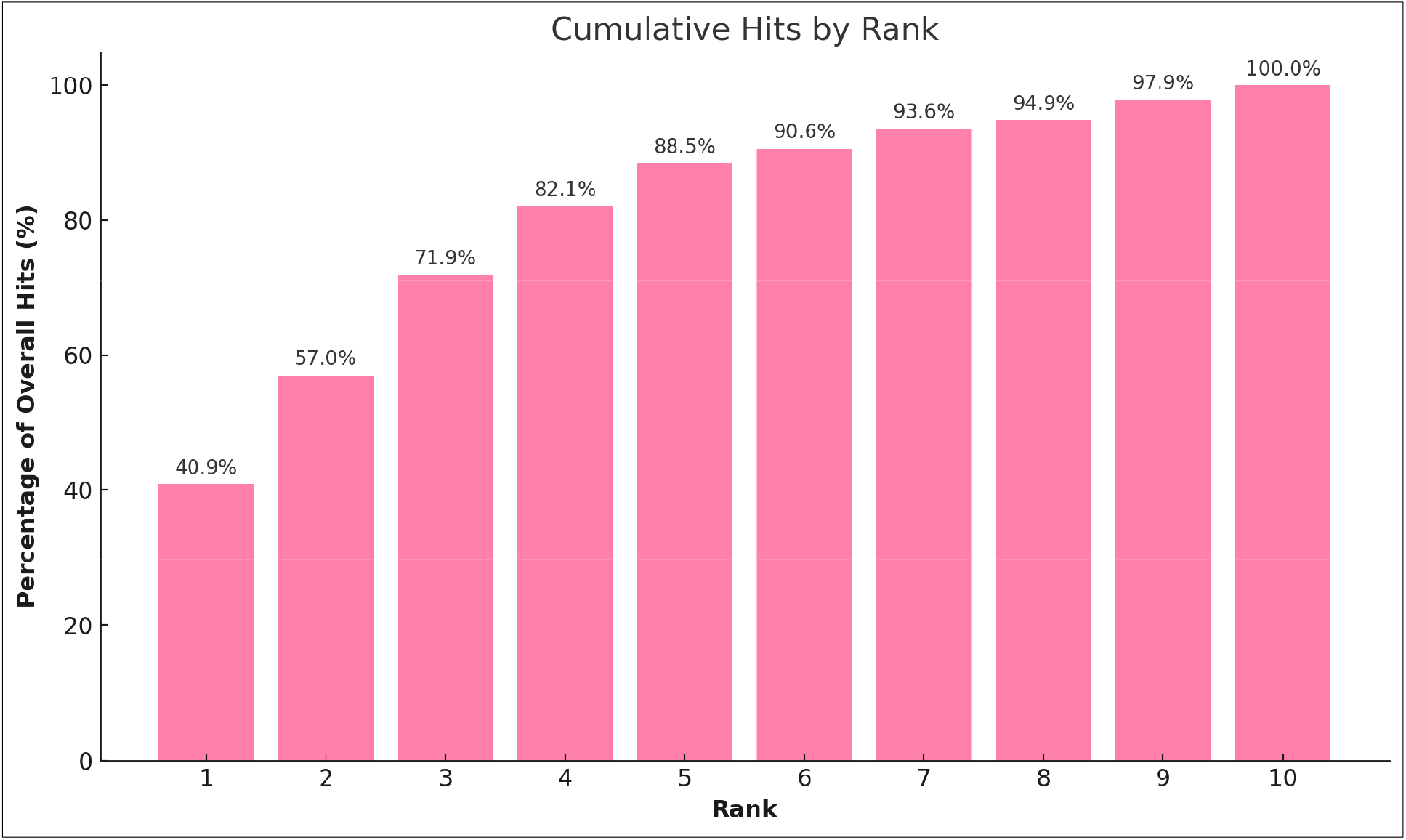
Cumulative Hits by Rank (Top 1 through 10)

While the overall Top 5 hit accuracy was 63.4%, there was a range of Top 5 hit accuracy when subset by ICD grouping from 0% to 86.7% (Figure 3). Categories that performed better than overall average included: nervous system, ear-related complaints, genitourinary system, obstetrics, respiratory system, endocrine system and digestive system. Categories that performed

**Figure 3.**
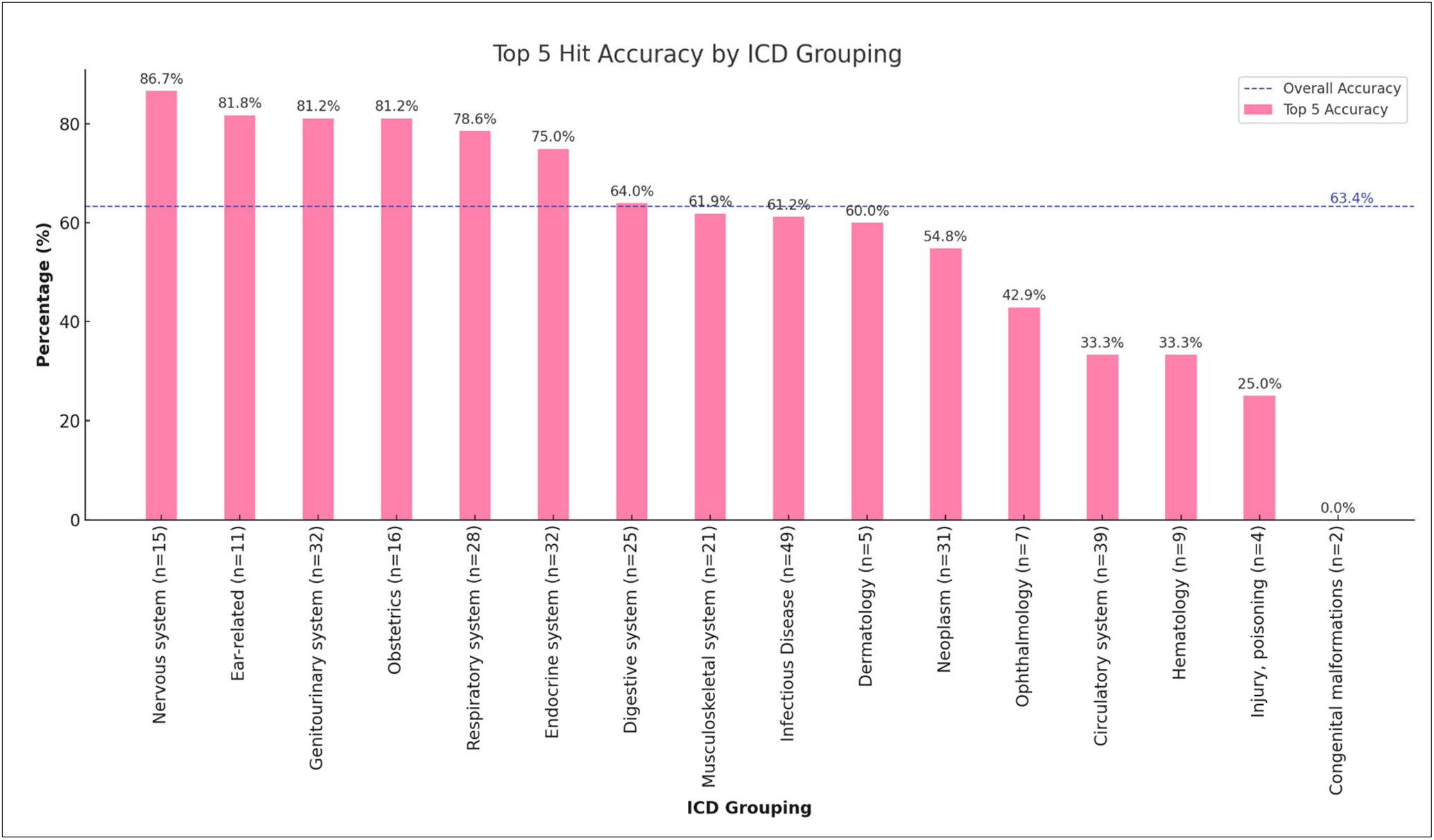
Top 5 Hit Accuracy by ICD Grouping.

**Figure 4.**
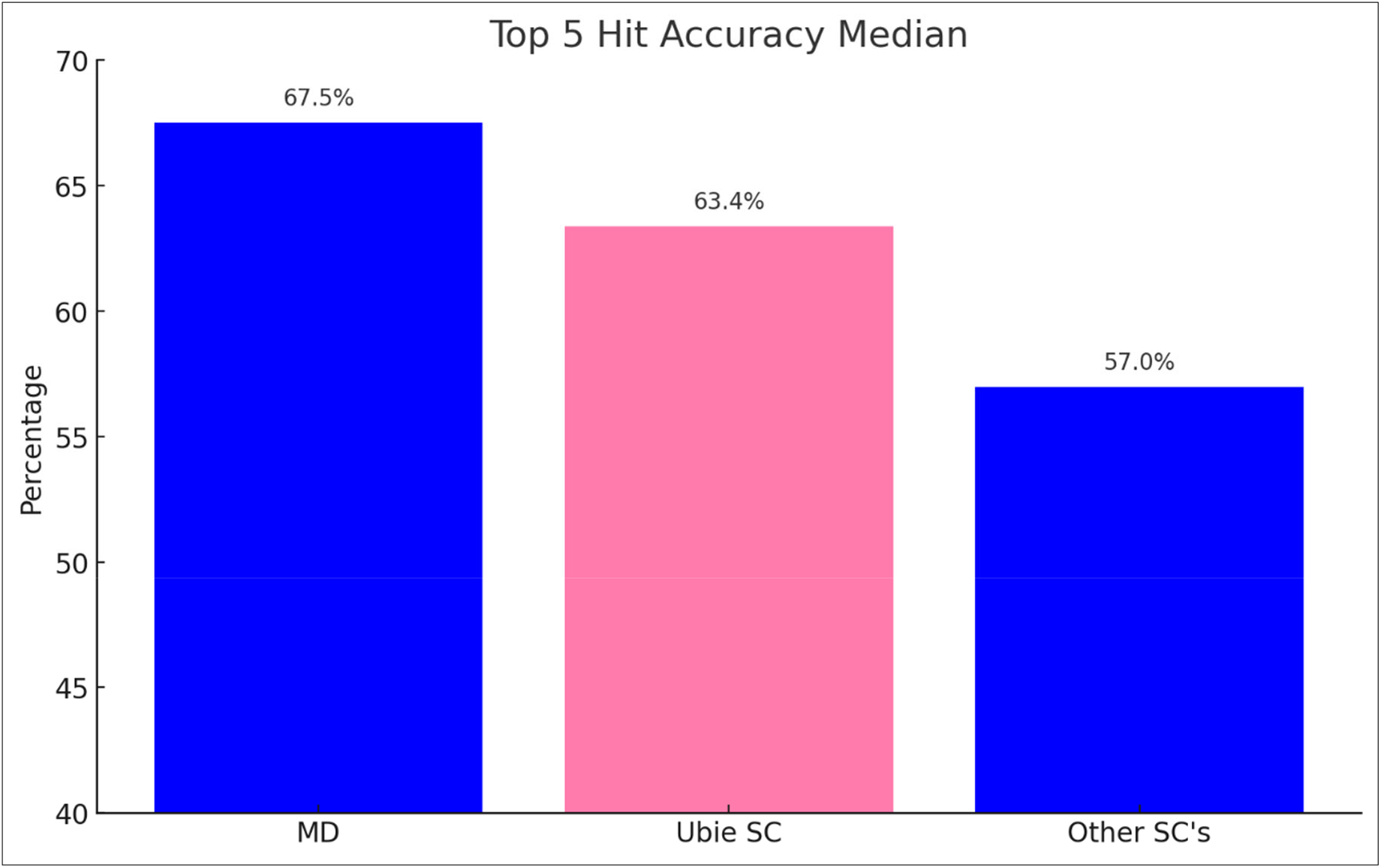
Top 5 Hit Accuracy Comparison.

### C. Comparison to Other SC’s and Physicians

Using publicly available data from the symptom accuracy of the study SC was compared to commercial SCs and physicians who also completed all 400 vignettes [11]. The performance comparison included median scores for MD-based checkers (MD1, MD2, MD3), a study-specific SC, and the median performance of below the overall average and with more than 5 vignettes included: musculoskeletal, infectious disease, dermatology, neoplasms, ophthalmology, circulatory system and hematology. Polycystic kidney disease and renal artery stenosis were the two vignettes that were classified in the ICD-10 system for congenital malformations.

5 other commercial SCs. Analysis revealed that the MD Average achieved the highest hit accuracy at 72.9% (62.7-88.5%). This average represents a combined metric for MD1, MD2, and MD3. The Study SC demonstrated lower accuracy of 63.4% but above the average for the other SC checkers, 57.0% (47.8-76.3%).

## Discussion

The results of this study highlight the study SC’s capabilities as an AI-driven tool for suggested diseases related to input symptoms, with a Top-5 hit accuracy of 63.4% and a Top-10 hit accuracy of 71.6%. These metrics indicate that the study SC is effective in providing relevant disease information within its top-ranked results, which can be a valuable asset in assisting patients with initial health assessments. The system’s performance across various clinical domains, particularly in areas such as the nervous system and respiratory conditions, suggests that Ubie is well-equipped to handle a diverse range of symptoms, some better than others.

The study successfully mapped almost 8000 data points with the aid of LLM’s and the process of medical entity linking. The study’s innovative use of AI-assisted simulation in evaluating Ubie represents a significant advancement over traditional static vignette studies, offering a dynamic and scalable method to assess the accuracy of symptom checkers. This approach addresses the common limitations of vignette studies, such as small sample sizes, static datasets and resource intensiveness, by enabling large-scale, automated simulations that can easily be re-simulated under different parameters to provide a more comprehensive understanding of a symptom checker’s performance. The inclusion of both present and absent symptoms in the simulation also enhances the clinical relevance of the study, as it mirrors the complexity of real-world diagnostic scenarios where both positive and negative findings contribute to clinical reasoning.

However, the variability in accuracy across different ICD groupings points to areas where further refinement is needed. For example, while Ubie SC performs well in certain domains, there is room for improvement in categories such as musculoskeletal and infectious diseases. Addressing these gaps will be crucial in enhancing the system’s overall effectiveness and ensuring it can provide reliable symptom-related information across a broader spectrum of conditions.

This study has several limitations. The use of simulated clinical vignettes, while useful, may not fully capture the complexities of real-world patient interactions, limiting the generalizability of the findings. The exclusion of vignettes outside the scope of the Ubie SC indicates gaps in the symptom checker’s capabilities. Additionally, the controlled testing environment does not replicate the dynamic nature of real-time symptom assessment, where user input and symptom evolution can significantly impact hit accuracy. For example, in the web-based SC, users can enter as many positive findings as they would like. Those positive responses in turn will determine how many negative responses they may enter.

Further research is needed to fully understand Ubie’s SC performance in more varied and real world clinical settings.

In conclusion, the Ubie SC shows considerable potential as an educational support tool in healthcare, particularly for patients seeking initial information on their symptoms. With reasonable accuracy, these patients can then be presented with relevant education and messaging based on listed related symptom-related disease that can guide them in future conversations with a health care provider. While there are areas that require ongoing development, the system’s strong performance in many clinical domains and this innovative evaluation method suggest AI SC’s may play a valuable role in the future of AI-assisted healthcare.

## Data Availability

All data produced in the present work are contained in the manuscript

## Notes

### Competing Interest Statement

NKT serves as a consultant for Ubie Inc., and TN is an employee of Ubie Inc.

### Funding Statement

This study was funded by Ubie Inc.

## Reference

1 Meyer, A. N., Giardina, T. D., Spitzmueller, C., Shahid, U., Scott, T. M., & Singh, H. (2020). Patient perspectives on the usefulness of an artificial intelligence-assisted symptom checker: Cross-sectional survey study. Journal of Medical Internet Research, 22(1), e14679.

2 Wallace, W., Chan, C., Chidambaram, S., Hanna, L., Iqbal, F. M., Acharya, A., Normahani, P., Ashrafian, H., Markar, S. R., Sounderajah, V., & Darzi, A. (2022). The diagnostic and triage accuracy of digital and online symptom checker tools: A systematic review. NPJ Digital Medicine, 5(1), 118. 10.1038/s41746-022-00667-w

3 Hill, M. G., Sim, M., & Mills, B. (2020). The quality of diagnosis and triage advice provided by free online symptom checkers and apps in Australia. The Medical Journal of Australia, 212(11), 514–519. 10.5694/mja2.50600

4 Semigran, H. L., Linder, J. A., Gidengil, C., & Mehrotra, A. (2015). Evaluation of symptom checkers for self-diagnosis and triage: Audit study. BMJ, 351, h3480. 10.1136/bmj.h3480

5 Painter, A., Hayhoe, B., Riboli-Sasco, E., & El-Osta, A. (2022). Online symptom checkers: Recommendations for a vignette-based clinical evaluation standard. Journal of Medical Internet Research, 24(10), e37408. 10.2196/37408

6 Shah, R., Edgar, D., & Evans, B. J. (2007). Measuring clinical practice. Ophthalmic and Physiological Optics, 27(2), 113–125. 10.1111/j.1475-1313.2006.00481.x

7 Kopka, M., Feufel, M. A., Berner, E. S., & Schmieding, M. L. (2023). How suitable are clinical vignettes for the evaluation of symptom checker apps? A test theoretical perspective. Digital Health, 9, 20552076231194929. 10.1177/20552076231194929

8 Hammoud, M., Douglas, S., Darmach, M., Alawneh, S., Sanyal, S., & Kanbour, Y. (2024). Evaluating the diagnostic performance of symptom checkers: Clinical vignette study. JMIR AI, 3, e46875. 10.2196/46875

9 Stoilos, G., Geleta, D., Shamdasani, J., & Khodadadi, M. (2018). A novel approach and practical algorithms for ontology integration. In Vrandečić, D., et al. (Eds.), The Semantic Web – ISWC 2018. ISWC 2018. Lecture Notes in Computer Science (Vol. 11136). Springer, Cham. 10.1007/978-3-030-00671-6_27

10 French, E., & McInnes, B. T. (2023). An overview of biomedical entity linking throughout the years. Journal of Biomedical Informatics, 137, 104252. 10.1016/j.jbi.2022.104252

11 Hammoud, M., Douglas, S., Darmach, M., Alawneh, S., Sanyal, S., & Kanbour, Y. (2024). Evaluating the diagnostic performance of symptom checkers: Clinical vignette study. JMIR AI, 3, e46875. 10.2196/46875

